# Acceptability and sustainability of a point-of-care HPV ‘self-collect, screen-and-treat’ for cervical cancer prevention in Papua New Guinea: A qualitative exploration of key informants’ perspectives

**DOI:** 10.1101/2023.09.27.23296249

**Authors:** Hawa Camara, Somu Nosi, Gloria Munnull, Steven G. Badman, John Bolgna, Joseph Kuk, Glen Mola, Rebecca Guy, Andrew J. Vallely, Angela Kelly-Hanku

## Abstract

**Introduction:** Innovative technologies over the past decade have emerged to increase uptake in cervical cancer early detection and treatment that could significantly improve screening and precancerous treatment. The changing landscape in cervical cancer screening algorithms and technologies calls for critical inquiries into their implementation in all settings, but especially in low-resource settings with the heaviest burden of disease. Papua New Guinea (Papua New Guinea) has among the highest estimated burden of cervical cancer globally yet has no organized national cervical screening programs. To better understand key informants’ perspectives of a same day point-of-care HPV screen-and-treat program, we conducted key informant interviews to capture their insights into the factors impacting the acceptability and sustainability of the intervention.

**Methods:** We conducted a total of 26 semi-structured interviews with a purposive sample of 20 health care workers and six policymakers from Well Woman Clinics in Madang (Madang Province) and Mt. Hagen (Western Highlands Province). Interviews were conducted in English, transcribed, and analysed using thematic analysis, highlighting factors impacting the acceptability and sustainability of the program from these key informants’ perspectives.

**Results:** The participants perceived the intervention as culturally fit and valuable. Health care workers agreed that the technological elements of the intervention were easy to use and provided the benefit of same day screen-and-treat, which helped to significantly reduce loss to follow-up. Factors such as planning for key resources (i.e., financing, and human resources) and political support were recognised as essential to ensure long-term sustainability by policymakers. The intervention was valued as ‘scalable, portable and simple’, emphasizing that key political support and a comprehensive national cervical cancer prevention strategy could help Papua New Guinea make considerable headway toward cervical cancer elimination.

**Conclusion:** In light of the burden of cervical cancer in the country, all participants agreed that a national cervical screening program, explicitly same day screen-and-treat services using self-collection, addressed an immense unmet need and salient cultural and systemic barriers.

**Contribution to literature:**

- This study is the first to explore factors impacting the acceptability and sustainability of a point-of-care HPV self-collect same day screen-and-treat in a low resource setting
- The intervention was positively welcomed and assessed by all key informants
- Both health care workers and policymakers agree that this service is culturally congruent, easy to use and implement, and is sustainable with consistent political and financial support
- Key socio-cultural and systemic factors were identified as caveats that, if raised, could potentially increase acceptability and sustainability of the program

## Introduction

The emergence of molecular-based testing platforms and treatment technologies have reshaped the early detection and treatment of cancers, most notably for cervical cancer, the fourth most common cancer among women [1]. In 2020, the World Health Organization (WHO) outlined a global strategy for the elimination of cervical cancer. Following HPV vaccination to prevent infection, the two remaining pillars are designed to ensure no women and others already infected with oncogenic HPV types die prematurely and needlessly from cervical cancer. In July 2021, WHO updated its guidelines to recommend HPV screen-and-treat as the primary screening modality [2]. As part of this impressive effort, and in recognition of the significant role it can play in low- and middle-income countries (LMICs), WHO now recommends same day human papillomavirus (HPV) screen-and-treat [3].

### Technological innovations for early detection and treatment of cervical changes

With the use of innovative technological platforms, it is now possible to detect oncogenic HPV types known to cause cervical cancer in cervical samples using a molecular platform using a Polymerase Chain Reaction (PCR)-based assay [4, 5]. This technology has higher sensitivity and specificity than conventional cytology methods for the detection of HPV, which requires women and others at risk of cervical cancer to undergo cervical screening less frequently, and some PCR tests are in formats that allow point-of-care by staff with minimal laboratory training [5–10]. The point-of-care feature is particularly important in LMICs due to women facing challenges accessing screening services associated with distance to health clinics, transportations costs (for repeated visits), and household responsibilities [11–13]. Moreover, with effective training, the technology allows for task-sharing among health care workers [14].

In settings where cervical screening uses traditional approaches such as the Pap test, the visual inspection with acetic acid (VIA) and even clinician-collected swabs for HPV screening, women’s participation in screening programs has been declining over the years. One of the reasons noted to be causing the decline is the embarrassment from the pelvic examination, among other deterrents [15, 16]. Persistent socio-religious and cultural beliefs have impacted women’s experience with and perceptions of the pelvic examination [17]. Research shows that sexual practices involving the female reproductive system, such as the Pap tests’ pelvic exam, engenders feelings of shame, embarrassment, and guilt, especially in countries with diverse cultural backgrounds such as Papua New Guinea (Papua New Guinea) [18–20]. To address the shame and stigma from the clinician-collected method (i.e., pelvic examination with the Pap test), scientists developed the self-sampling (or self-collection) device that allows for women to collect their own cervical samples. This novel collection method has shown to be comparable to clinical-collected samples [9, 21, 22] and is widely preferred by women and others [23–26], citing its private nature, ease of use, and increased access to the health system. Self-collection is increasingly being used across the world in cervical screening programs, enabling women to self-collect in the clinic or even in the privacy of their own home and post the sample back to the laboratory [27–29]. Self-collection has the potential to significantly increase participation in screening programs in all settings, most notably low-resource settings [25, 30–34].

### Cervical screening in Papua New Guinea

In Papua New Guinea (Papua New Guinea), cervical cancer is the second most common cancer [35]. In 2020 alone, Papua New Guinea recorded an age-standardized (ASR) cervical cancer incidence rate and mortality rate of 29.2 cases per 100,000 and 19.1 cases per 100,000 women, respectively [36]. Past screening services were reliant on Pap tests. This now outdated screening method was reliant on highly trained staff, was resource intensive and without adequate laboratory facilities could not be taken to scale. With cytology processing occurring offshore, in Australia, the turnaround times meant women went without receiving their results. In 2009, a pilot study was conducted in Papua New Guinea which used Visual Inspection with Acetic Acid (VIA) and ablative cryotherapy [10] to provide same-day testing and treatment. However, the use of VIA had low sensitivity and specificity which led to misdiagnoses and overtreatment and required a pelvic/clinical examination. VIA also failed to overcome the barrier of an examination.

In 2018, a field prospective single-arm intervention study of same day screen/test-and-treat HPV-based screening (HPV ‘self-collect, test and treat’ or HPV STAT) using self-collect, POC testing and thermal ablation was implemented in two provinces in Papua New Guinea [5, 10, 37]. The testing platform selected for this prospective single-arm intervention study was the Xpert (GeneXpert) HPV test, which has been used in numerous LMICs in the diagnosis of infectious diseases such as tuberculosis and other sexually transmitted infections [38]. The Xpert (GeneXpert) HPV test allows for 60-90 minutes turnaround time for results, compared to a few hours for other similar platforms. In performance studies, Xpert has shown to be an easy-to-use, scalable, effective, and reliable diagnostic technology for point-of-care HPV testing [4, 5, 39, 40]. The clinical algorithm used is detailed elsewhere [37].

### Key informants and cervical screening

Health workers, along with program managers, are highly respected for their in-depth understanding of the community, culture, and their role in the implementation of health interventions [41, 42], while policymakers understand the health system and can offer their own perspectives about systemic factors that could impact program implementation. Exploring the perspective and experiences of key informants such as health care workers can offer valuable insights into how socio-cultural, clinical, and systemic factors collide, to help catalyse investments in cervical cancer prevention and control nationally [43].

Based on earlier social research in Papua New Guinea which sought to contextualise the socio-cultural and biomedical understanding of cervical cancer and HPV vaccination for prevention showed that women’s understanding of what cervical cancer is, and what causes the disease was ambiguous at best [44]. This same research showed that although knowledge of HPV as the causative agent for cervical cancer was poorly understood, once the link was made, there was widespread support from women, men, and key informants for a national roll out of HPV vaccination; where there was discrepancy was between older and younger people about the heath communication messaging of the purpose of the vaccine [45, 46]. More recent qualitative work has shown that women who have participated in Papua New Guinea’s HPV STAT that self-collection was highly acceptable. As part of this same HPV STAT program researchers have shown that the PCR platform used for testing is efficacious and effective [5, 10, 47]. The next piece of this growing body of knowledge about self-collected HPV screen-and-treat is the experiences and perspectives of health workers and policymakers of a point-of-care early detection and treatment cervical cancer program in the country.

As approaches to cervical screening rapidly evolve and are trialled and implemented in diverse geographical as well as socio-political contexts, and global efforts are scaled up to eliminate cervical cancer, it is imperative to continue to evaluate such programs from the point of view not just of women but assess programs from the perspective of key informants. Through this study and the data presented here in this paper we start to address this gap, examining HPV STAT from a social research and implementation research perspective in an attempt to inform national policies for cervical screening and treatment services in a high burden setting. This qualitative study explores perspectives and experiences of key informants to understand the acceptability and sustainability of a self-collected same day HPV screen-and-treat program in Papua New Guinea, a setting where new approaches are urgently needed.

## Materials and Methods

A qualitative study was undertaken as part of the larger multidisciplinary study exploring the performance, cost-effectiveness, acceptability, and health systems requirement of a same day approach for early detection and treatment of cervical precancerous lesions, using self-collection. The field prospective single-arm intervention study was conducted at two Well Women Clinics (WWC) in provincial hospitals in Papua New Guinea (Modilon General Hospital, Madang Province and Mount Hagen General Hospital, Western Highlands Province). On arrival at the clinics all attending women are given a health education talk (“Tok Save”) by health workers. The health education talk covers risks and causes of cervical cancer (i.e., HPV as a cancer-causing sexually transmitted infection), instructions for self-collection, and information about the same screen-and-treat service, including how women who test positive for high-risk HPV (hr-HPV) will be treated and triaged. All health care workers were trained to operate the testing platform and interpret the results.

### Study population and setting

Semi-structured interviews were conducted with 26 key informants (20 health care workers and 6 policymakers) in Madang and Western Highlands Provinces. All interviews took place between June 3^rd^ and August 16^th^, 2019. We used purposive sampling to identify key health care workers and policymakers with institutional knowledge and direct or indirect experience of implementing HPV STAT [48]. The key informants were identified with the help of the PNGIMR team, and the prospective single-arm intervention study’s principal investigator. During fieldwork, the pre-identified key informants were called, emailed, or directly approached and asked to participate in the interviews. This ensured a heterogeneous sample of participants with insightful, diverse, and information rich narratives to address the research question at hand. Health care workers included medical/nursing directors, senior hospital health care workers members, including nurses, Obstetrics and Gynaecology ward staff and health extension officers (HEO). Policymakers included senior-level directors or managers with over 15 years of experience in the national health system. All policymakers involved were part of the Provincial Health Authority and were key decisionmakers in health policy in their province.

### Data collection and analysis

The interviews ranged in length from 30-80 minutes (mean: 45 minutes). Two researchers (HC and SN) conducted the interviews in English, and all were digitally audio-recorded. The first and second authors conducted the interviews, with the second author being an experienced social researcher from Papua New Guinea fluent in both English and Tok Pisin. With the help of the Papua New Guinea Institute of Medical Research (PNGIMR), staff helped transcribe and translate all the Tok Pisin interviews, while the first author transcribed all the interviews conducted in English.

All participants were given an information and consent form in either English or Tok Pisin. The form was explained to each of the participants to ensure all participants understood the purpose of the interview, provided consent to participate, and signed the consent form before starting. The interviews took place in conference rooms, office spaces, or at the clinic, ensuring the key informants’ privacy and security. All participants have been anonymized to protect their identities. We used pseudonyms for all quotes in this paper to ensure our participants’ anonymity.

The first author used an interpretive approach to the data analysis. The process was iterative throughout the analysis to capture experiences and perspectives impacting HPV STAT’s acceptability and sustainability. The first author conducted a line-by-line inductive thematic analysis outlined by Braun and Clarke [49] to analyse the transcripts using the following six steps: (1) transcription, (2) reading and re-reading to become familiar with the data, (3) coding, (3) search for themes, (4) review the themes, (5) define and name the themes, and (6) finalize the analysis. The lead author coded each transcript independently using QSR NVivo 12.5 [50]. The first author focused on ‘micro to macro’ factors impacting acceptability and sustainability of the program. Accordingly, she identified sub-themes that characterized their perspectives and experiences impacting acceptability and sustainability from a personal, an interpersonal, and from a health systems/policy point of view. These sub-themes were grouped into themes that spoke to a social, clinical, and systemic views of the program. The author provided illustrative quotes for each theme/sub-theme.

### Ethics

The team received ethical approval for this study and the larger prospective single-arm intervention study from the Papua New Guinea Institute of Medical Research (PNGIMR) Institutional Review Board (IRB), the Medical Research Advisory Committee (MRAC) of the Papua New Guinea National Department of Health (NDoH), and the Human Research Ethics Committee at UNSW Sydney.

## Results

### Participant demographics

Of the 26 key informants interviewed, the majority were working and living in Madang (n=17, 65%). The key informants held several different types of positions in the provincial health service including program directors (n=6, 23%), specialist Obstetricians/ Gynaecologists (n=5, 19%) and nurses (n=5, 19%). Reflective of the health workforce more generally [51], most were female (n=21, 81%). Of all the participants, most (n=17, 65%) had more than 15 years of experience in Papua New Guinea’s health system, with an average of 16.5 years.

**Table 1.** Participants demographics per site.

| Categories | Sub-categories | Madang |  | Mount Hagen |  | Combined |  |
| --- | --- | --- | --- | --- | --- | --- | --- |
|  |  | N=17 | 65% | N=9 | 35% | N=26 | 100% |
| **Role** | *CHW* | 2 | 12 | 1 | 11 | 3 | 12 |
|  | *Director* | 1 | 6 | 5 | 56 | 6 | 23 |
|  | *Health Manager* | 0 | 0 | 1 | 11 | 1 | 4 |
|  | *HEO* | 4 | 24 | 0 | 0 | 4 | 15 |
|  | *Midwife* | 2 | 12 | 0 | 0 | 2 | 8 |
|  | *Nurse* | 3 | 18 | 2 | 22 | 5 | 19 |
|  | *Physicians (Ob/Gyn)* | 5 | 29 | 0 | 0 | 5 | 19 |
| **Sex** | *Female* | 16 | 94 | 5 | 56 | 21 | 81 |
|  | *Male* | 1 | 6 | 4 | 44 | 5 | 19 |
| **Years in Papua New Guinea Health System** | *Less than 5 years* | 4 | 24 | 0 | 0 | 4 | 15 |
|  | *5-10 years* | 1 | 6 | 0 | 0 | 1 | 4 |
|  | *10- 15 years* | 4 | 24 | 0 | 0 | 4 | 15 |
|  | *More than 15 years* | 8 | 47 | 9 | 100 | 17 | 65 |

## Findings

Based on a thematic analysis there were three main overarching themes that were central to the acceptability of the screening intervention: (1) embracing HPV STAT’s impact, values, and opportunities: a social perspective; (2) engaging with point-of-care technology: a clinical perspective; and (3) ensuring sustainability: a systems perspective. Themes 1 and 2 explore factors influencing key informants’ acceptability of HPV STAT, and theme 3 explores the salient factors required to sustain the program.

**Diagram 1.**
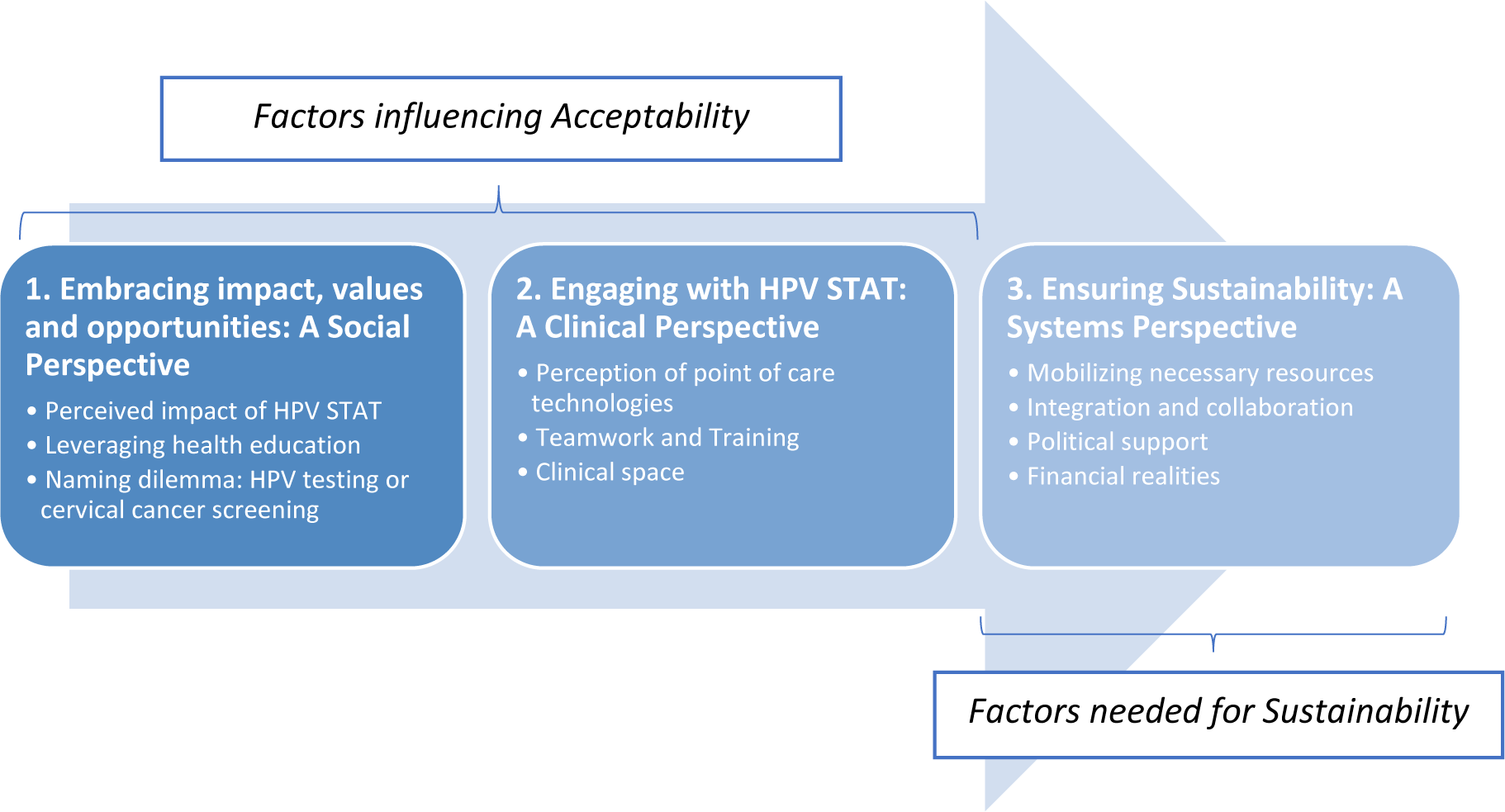
Thematic Analysis identifying factors impacting acceptability and sustainability

### Embracing HPV STAT’s impact, values, opportunities: a social perspective

The theme of ‘embracing HPV STAT’s impact, values, and opportunities’ details how the participants perceived the intervention from a social perspective. In this section, we review key stakeholder’s recognition that the program addresses key socio-cultural barriers and provides opportunities for a wider national impact.

#### Perceived impact of HPV STAT

A key discussion point in the interviews was cervical cancer’s insidious nature. The lack of accessible and culturally congruent prevention services meant that women were dying early, and most often experienced a physically and emotionally painful period of isolation in the later stages of the disease due to the lack of treatment options. All key informants highlighted the imminent need for effective and culturally sensitive preventive services against cervical precancer and cancer. They perceived HPV STAT (‘the program’) as a social and clinical necessity to reduce the burden of disease nationally. They emphasized that the program, as designed, addresses numerous socio-cultural and systemic barriers that have hindered women from participating in previous programs.

Another major cultural impact of the program is the self-collection of vaginal swabs, which was identified as having a significant impact on women’s willingness to participate in screening. Earlier strategies based on Pap test collection or VIA examination required all women to undergo a pelvic examination which are poorly accepted due to embarrassment, discomfort, and cultural contexts.

> When we did the VIA [visual inspection with acetic acid], the mothers got scared like they were ashamed because of our customs and all these things. They wouldn’t just come and open themselves up for anybody to see them; they were a bit ashamed to come. But today when we are saying that you will do your self-collection, a lot of mothers are coming up to do that. They feel more comfortable in that way. (Marjorie, health care worker, 22 years of experience)

With the availability of self-collection, women have become even more active in their own care, while at the same time becoming more self-aware of their bodies.

> I see that [self-collection] has really encouraged women to come forward because they are getting the specimen by themselves which they feel more comfortable and also, we respect their privacy. They are empowered and they take ownership of their own health, of their own well-being. I see that, that’s very good, excellent idea because like I said previously, they don’t want to show themselves to other people. (Eloise, health care worker, 20 years of experience)

In previous Pap test programs, several barriers contributed to most women being lost to follow-up: the financial burden of transportation costs for frequent travel to the clinic (i.e., visit for the screening, and return for the results and ongoing care), as well as delays in receiving their Pap test results. The single visit (same day) point-of-care strategy allows for women to be tested and treated on the same day, greatly reducing loss to follow-up, opportunity costs, and promoting participation in screening. Senior health care workers and policymakers recognised that the women-led program provides the directions, guidelines on how the country should move forward in managing the disease.

> Cervical cancer is becoming more common. This is an emerging disease that is killing a lot of women in Papua New Guinea. So, when you think about something that could actually save young girls and women from dying of a very high-risk disease… that is simple, portable, scalable, women-led… we like it! (Jaden, policymaker, 20 years of experience)

#### Leveraging health education

These one-hour sessions create an environment where women become more fully informed about the service. This was an opportunity for health care workers to address individual concerns and misunderstandings of/about the screen-and-treat procedure, such as seeking clarification about the self-collection procedure, timing of results, and how to prevent cervical cancer in the future.

A few caveats were raised by health care workers around the introduction of HPV as a causative agent for cervical cancer. Despite attending the health education sessions, a few women continued to believe that a positive HPV test meant they had cervical cancer. When discussing this issue in their interviews, some health care workers stressed the importance of clarifying the difference between testing positive for HPV and being diagnosed with cervical cancer to avoid any misunderstandings and conflation of the two.

> At first when we started doing it, those who tested positive were saying ‘I have cancer’, and they think that we can’t kill that cancer, but when they come back, we said, ‘No, this is not cancer.’ The women who came after, we explain to them that if you have that virus, it doesn’t mean that you have cervical cancer. Just a virus is there, you don’t necessarily have cervical cancer. (Jaylene, health care worker, 13 years of experience)

Having received thermal ablation to treat the precancerous lesions, women who had tested positive for HPV and undergone same day treatment were advised to abstain from vaginal intercourse for six weeks. Abstinence is recommended to minimize the side effects of the thermal ablation and promote cervical healing. According to key informants, for some of the women, this post-care requirement created mistrust with their partners/husbands. To date men have not attended the Well Women’s Clinic and have therefore not been privy to the health education talks that explain this medical necessity for abstinence. The underlying reason for needing to abstain –and to promote the healing process – is the issue that causes most of the tension in relationships.

#### A naming dilemma: HPV testing or cervical cancer screening?

Both health care workers and policymakers raised the importance of finding the correct way to ‘name’ or ‘describe’ the service and viewed it as an opportunity to reduce the stigma around HPV and cervical screening. In Papua New Guinea, discussions around topics such as sex, sexually transmitted infections (STI), or the reproductive system can be highly stigmatized. Thus, naming the testing service provided is of cultural significance and would profoundly impact acceptability and uptake. The majority of health care workers and policymakers believed that the testing (should be referred to as ‘cervical screening’ or ‘HPV testing’, albeit they acknowledged that calling it HPV testing could be considered an important step in the right direction and would help lessen any dissonance in communication pertaining to the service.

> Using HPV testing is bold because people have to know that HPV causes cervical cancer. It’s like people are going for HIV testing. Now, we’ll have to say that HPV testing is available too. (Janelle, health care worker, 2 years of experience)

While the use of ‘HPV testing’ may be considered ‘bold’, some cautioned that it is important to be sensitive and responsive to the stigma associated with HPV being an STI and to the diverse cultural realities of each province in the country.

> We need to be sensitive that what works in one province, may need tweaking for it to be acceptable somewhere else. As long as we stick to the core things of it being something that is going to avoid deaths from cervical cancer, it is a hard message to argue with. Now, how you might talk about that STD part of HPV, to me, would be to dilute it, because the other message [preventing deaths] is so incredibly powerful. (Mayna, policymaker, 23 years of experience)

### Engaging with point-of-care: a clinical perspective

A key factor in evaluating the implementation of an intervention is assessing how it is viewed from a clinical perspective. In this section, we review the clinical intervention, the clinical space, and the clinical engagement.

#### Perceptions of HPV STAT’s point-of-care technologies

As part of the program, health care workers operated a diagnostic point-of-care technology to test for the presence of hr-HPV DNA. Trust in the diagnostic technology is apparent from discussions with both health care workers and policymakers. All health care workers were confident in the use of the GeneXpert platform including its accuracy. Most felt that the use of this closed molecular platform was more accurate than previous screening methods such as VIA.

Health care workers also acknowledged the quick result turnaround time of results using the GeneXpert, noting that it allows for same day screen-and-treat. Most health care workers noted the key aspects of time and speed, which help to address the high loss to follow-up rates experienced with past Pap test programs.

All health care workers who have operated the HPV testing platform said that it was easy to use. The testing platform interprets the results for the user, provides the exact HPV type(s) detected in the sample, and also indicates when there is an issue with the sample (i.e., not enough sample in the cartridge prompting for more sample collection). Similarly, policymakers agreed that the machine is ‘very effective and efficient’. Policymakers also commented on the machine’s mobility, alluding that it can be transported into hard-to-reach (rural) areas, where access to cervical screening remains a challenge.

The addition of the precancerous treatment option on the same day was seen as an advantage. All health care workers viewed the thermal ablation procedure as quick and effective.

> …with this [intervention], they come in and then they get the results on the same day and it’s possible they get treatment on the same day as well. That [same day screen-and-treat] would be our greatest benefit to the women. (Jeffrey, health care worker, 16 years of experience)

#### Teamwork and Training

Staff at both WWC saw their role and the role of others in their team as equally important. The effective teamwork stemmed from the clear delineation of roles as well as task sharing. The ’test and treat’ technologies are easy to use, allowing less skilled staff to operationalize Xpert, while also leading health education sessions, and counselling the women throughout the clinical pathway. The health care workers took particular pride in providing post-testing counselling, seeing it as a significant role in the women’s experience of HPV STAT.

The training model is one of coaching and supervision. Training consists of newly hired staff, namely health extension officers and nurses, observing the most experienced health care workers and being coached to perform all the steps. Task-shifting is an important advantage of this intervention as all health care workers directly involved are health extension officers and/or nurses, emphasizing the importance of their role in the intervention and in the Papua New Guinean health system. The coaching and supervision scheme promotes a mentoring environment. The staff felt supported by one another, creating a trusting work environment. The staff felt empowered, not only through the use of the Xpert platforms but the role they each led and being viewed as champions in their clinical and social environments. Policymakers suggested that a similar coaching strategy should be used during scale-up to empower clinic staff, to optimize on resources, and to create a supportive environment for all team members.

> It seems to me that one of the biggest reasons this is going really well is the passion and engagement [staff members] bring to our program. That’s going to mean that we don’t just take our staff out of here, but we’ve got to grow someone like them in other places and bring that person here for training, and then do the same thing again somewhere else. (Mayna, policymaker, 23 years of experience)

#### Clinical Space

According to all health care workers, the clinical space was well organized. Standard operating procedures and other key documentation were available and well within reach of the staff. The clinical algorithm posters were helpful to refer to when having conversations with the women regarding the intervention. Nonetheless, the majority of health care workers suggested improvements in a few areas. The size of the clinic was deemed too small. Due to the high uptake, there was no adequate space for the women to sit and wait for their results. At times, the women were too close to one another and could hear the other’s results during their post-test counselling. As much as the health care workers would attempt to ensure confidentiality during those sessions, the limited space made it challenging.

> These women are just sitting close to each other… we are sitting too close to interview clients, and we need to keep their privacy. So, they’re sitting close, and the others are listening on the other side, it is not good. When we give results, others are still listening. They’ll go back and say this woman has that virus or something. We need to keep things confidential. (Jaylene, health care worker, 13 years of experience)

As part of the program and to ensure women’s privacy, a private space was dedicated to the self-collection procedure and for women needing thermal ablation. Health care workers also acknowledged the value of the dedicated private space for self-collection and the thermal ablation procedure as part of the program. Similarly to the post-result counselling sessions, health care workers emphasised the need for a larger clinical space in order to accommodate the women and ensure privacy for these procedures. Moreover, the limited availability of a cervical screening program in Papua New Guinea has left a void that HPV STAT is filling. Most health care workers predicted an uptake due to increased awareness (i.e., word of mouth), high acceptability, and immense need. It was suggested that, when considering the impact the service could have on Papua New Guinea women long term, the service should have a space of their own to respond to the demand.

> If the hospitals all were willing to sustain the services, I think they should have a building of its own so there’s enough space, because it’s a service in demand for women in the province. (Jenny, health care worker, 17 years of experience)

### Ensuring Sustainability: a systems perspective

A systems perspective focuses on how the intervention interacts with and impacts on the system as a whole. The following will highlight key areas of improvement and/or consideration for scale up and sustainability.

#### Mobilizing necessary resources

Due to high acceptability of the program, health care workers underscored the need to hire for more staff to sustain the service. Some of the health care workers suggested to train nurses, health extension officers, and midwives currently working at the hospital, as a short-term remedy.

> The staff ratio to the number of women is overwhelming. For this particular clinic, even with the two nurses that we have at the clinic, we are trying to strategize with the numbers, but we are still overwhelmed with the numbers. The nurses to patient ratio is still an extremely big gap. We can’t match it. We might need additional help from other departments as a start. (Jeffrey, health care worker, 16 years of experience)

#### Political support

The introduction of HPV STAT prompted discussions about the need for a comprehensive national cervical cancer prevention strategy including all preventive measures (i.e., HPV vaccination, cervical screening, cervical cancer treatment services) to be made available to the public. Policymakers stressed the importance of supporting the implementation of all three pillars of the WHO cervical cancer elimination strategy. Yet, in order to see significant gains in cervical cancer elimination nationally, it will require all pillars for sustained improved health outcomes in the long term. Actualising this requires strong political support, notably political will, and direction. This will ensure oversight of the infrastructure and logistics required to sustain the program, such as the purchase of testing and treatment equipment, and key consumables (i.e., cartridges).

> We will have to build that into the annual plan. There are some definite plans in place, but that’s for the hospital. We have to take it right across the province then. We need good political will and directions from people higher up. They can get involved in the infrastructure and logistics side of it and buy the machines and sustain the service with getting its consumables like the cartridges and things like that. (Carl, senior policymaker, 22 years of experience)

Some respondents advocated for leadership at the provincial or national level to ensure more coordinated effort to sustain HPV STAT.

> The program needs a full-time person to coordinate and develop this. Bring everybody together and run a full cervical cancer screening clinic for the province as a whole. Everyone is busy with all the other responsibilities. So, to really scale it up across the province, that would be really nice if we can do it… even nationally. With good political direction and will, well-coordinated and a focal person, we can lead and sustain the program. (Roger, senior health care worker, 14 years of experience)

#### Financing realities

The cost and affordability of HPV STAT were raised by both health care workers and policymakers. The current service is provided free of charge. While working women may be in a financial position to pay in full or at least cost share as the program is taken to scale, it was recognized that for the majority of women in Papua New Guinea this would not be possible. Yet, both health care workers and policymakers recognized that it is essential to charge for the service if it needs to be sustained. Senior-level health care workers and policymakers emphasized that an effective financial strategy would allow the purchasing and maintenance of supplies and (extra) GeneXpert machines essential to respond to the demand generated with the introduction of the service.

> To really have an effective and sustainable program, women will have to be charged. I would think that 60 Kina would be sufficient. 50 Kina is for cartridge. That extra 10 Kina will be for treatment and many unseen costs that would be incurred. But that’s from the top of my head. I don’t want it to be so hard for the families. But at the same time, I don’t want to be so cheap, and we struggle and close down the clinic. So, this could be a balance. And if we have a reasonable balance like that, we ask the local MPs or the government to support it, and we can just try it that way to start. (Peter, policymaker, 25 years of experience)

## Discussion

On July 6^th^, 2021, the WHO launched its new guidelines for cervical cancer early detection and treatment strongly recommending the screen (or test) and treat approach used and being evaluated in Papua New Guinea since 2018. These findings on key informants’ acceptability of HPV STAT are thus, timely.

This is the first study to examine and report on the experiences and perspectives of health care workers and policymakers of a point-of-care early detection and treatment cervical cancer program in Papua New Guinea, and, as we understand, globally.

As part of our interest to explore factors impacting the implementation of a self-collect HPV screen-and-treat program, we considered health care workers and policymakers’ perceptions and experiences of HPV STAT, focusing on perceptions of acceptability (i.e., the perception among informants that a given treatment, service, practice, or innovation is agreeable, palatable or satisfactory) and sustainability (i.e., the extent to which an intervention is maintained or institutionalized in a given setting) [52]. The study has identified several important strengths of the HPV STAT intervention that impacted its acceptability and sustainability.

Using the innovative HPV testing at point-of-care enabled a continuum of care that was not possible in earlier cervical screening models, thus mitigating the unacceptably high loss to follow-up seen in earlier programs. In this paper we have shown that key informants (and elsewhere the women) trusted the PCR-based platform and its results and allowed for same day screen-and-treat, which was highly valued by all informants, and for those operating the testing machine, have found it was easy to use. Both health care workers and policymakers emphasized the value of the intervention and its potential impact on women in the program but also the community more widely. This is in line with the limited research that features the strengths of HPV testing platforms [53]. The use of self-collection augmented the positive perspectives of the intervention. Most health care workers perceived self-collection as private, easy to use and less invasive than the pelvic examination. This is consistent with previous research highlighting key features of the self-collection option [24, 26, 27].

These elements all contributed to the high acceptability of HPV STAT as a cervical screening intervention in Papua New Guinea. As with all new interventions, we were able to identify a few areas that should be considered during the scale-up of the program. Health care workers and policymakers were both adamant that ensuring proper human resources for health (HRH) coverage for the service is essential to sustain it. It was explained that lack of sufficient health care workers might lead to drop-offs in uptake and threaten staff retention. Some suggested strategies reinforcing task sharing with current staff at the hospital to address the imminent need. Previous studies in similar low-resource settings have shed light on these concerns [54–56]. However, increasing staff and potential integration may lead to additional financial burden. Sustaining the service, which include increasing staff, purchasing, and maintaining equipment and supplies, requires a financing strategy that will allow to cover recurring costs. In addition, integrating the service with other testing services (e.g., HIV testing and counselling) means potentially stigmatizing the HPV testing program. A recently published study, the first of its kind, detailed the costs and cost-effectiveness associated with HPV STAT. It was found that, HPV STAT, more specifically the 5-year screening strategy, is more effective and cost-effective over time compared to previous screening methods [57]. The study demonstrated that the same day screen-and-treat modality is the best ‘value for money’, especially in high burden settings with high loss to follow-up. The model predicts the annual cost of HPV STAT over the first five years at approximately 3.3 million USD. Additional research is needed to explore key hidden costs and potential solutions (e.g., costing, communication strategies) in Papua New Guinea and a myriad of high burden settings, to strengthen the evidence base.

There is a paucity of qualitative research focusing on same day HPV screen and treat. Most of the existing literature regarding same day ‘screen-and-treat’ pertains to the use of VIA as the screening method of choice [58, 59]. Further research is necessary to qualitatively explore the implementation of same-day screen-and-treat coupling point-of-care PCR-based technologies and self-collection. This will allow to gage preferences from diverse perspectives, including advantages and disadvantages of each option.

### Limitations and strength of the study

There are several limitations to this study. The use of a theory (e.g., CFIR, PRISM, RE-AIM) at the onset of the qualitative study could have helped strengthened the analysis of the results to cover difference aspects of acceptability and sustainability that were not highlighted in this manuscript. Including national-level policymakers would have enriched the findings to better inform the national strategy for cervical cancer prevention and control. Due to scheduling conflicts, one interview was conducted with two policymakers, one more senior than the other. This could have created some bias in the less senior respondent’s responses during the interview process. This study could have also included the women’s viewpoints to supplement the perspectives highlighted in the findings. The women’s perspectives, however, will be published in a separate article [60]. Because some of the health care workers’ interviews occurred after the interviews with policymakers, some of the health care workers’ findings could not be followed up on or discussed in more depth with policymakers about how the PNG health system could respond to caveats that were raised. Yet, these findings provide a foundation for further studies to confirm or add to the PNG experience.

The use of qualitative methods, however, presents a strength of the study. The use of interviews helped to explore in depth a research area that has not yet been qualitatively investigated. This study adds to the evidence base for an important public health measure, assessing two key implementation outcome measures in a high burden setting. The sample includes both health care workers and policymakers of different backgrounds and lengths of experience in the Papua New Guinea health system, offering a variety of perspectives that helped enrich the results. Yet, future research should include multiple other perspectives from other settings in the country for a more comprehensive exploration of socio-cultural, clinical, and systemic factors impacting acceptability and sustainability of HPV STAT. Moreover, additional qualitative research is needed in different high burden settings to complement the results presented in this study and add to the literature in support of the WHO screen-and-treat recommendation.

## Conclusion

The quote ‘simple, portable, scalable, women-led… we like it!’ summarizes the health care workers and policymakers’ perspectives of HPV STAT. This study highlights that innovative technologies in cervical cancer early detection and treatment, such as HPV STAT, can be sustained when key social, clinical, and systemic factors are considered during the design of the intervention. In light of the burden of cervical cancer in Papua New Guinea, this qualitative study demonstrated that a national culturally and financially appropriate cervical screening program, explicitly self-collected HPV-based same day screen-and-treat services, fostered engagement of health care workers and support from key policymakers. Whilst this study has raised a few caveats, it has also shown that HPV STAT presents opportunities that could be the answer to Papua New Guinea’s journey toward cervical cancer elimination.

## Data Availability

The data used and analyzed for this study were not made publicly available to protect participants’ privacy but are available from the corresponding author on reasonable request.

## Declarations

### Ethics approval and consent to participate

This study was conducted as part of a National Health and Medical Research Council (NHMRC) funded project grant (GNT1104938) ‘Prospective cohort study to evaluate point-of care HPV-DNA testing for the early detection and treatment of cervical pre-cancer in high-burden, low-resource settings.’ This study was approved by the PNG Institute of Medical Research Institutional Review Board (IRB) (No. 1712), the Medical Research Advisory Committee (MRAC) of the PNG National Department of Health (NDoH) (No. 17.36), and the Human Research Ethics Committee at UNSW Sydney (No. HC17631). All participants signed an informed consent before participating in the interviews.

### Consent for publication

Not applicable.

#### Funding

Funding was provided by the University of New South Wales scholarship Scientia scheme (to the first author). This study is part of a doctoral research thesis.

### Authors’ contributions

HC designed, collected, and analysed the data, wrote, and revised the manuscript. HC conducted and transcribed the interviews conducted in English. The Tok Pisin interviews were conducted, translated, and transcribed by SN. SN assisted with participant recruitment. AV, RG and AKH contributed to the design of the manuscript, the analysis, and the writing of the manuscript. All authors read, critically reviewed, and approved the final manuscript.

## Acknowledgements

The authors would like to thank all the participants, the PNG IMR team, Modilon Hospital Well Women’s Clinic and Mount Hagen General Hospital Well Women’s Clinic staff members for their instrumental support during the first author’s fieldwork. The authors would also with to acknowledge the senior level staff at both Modilon General Hospital and Mount Hagen General Hospital for their hospitality, time, and support of this study.

